# An analysis of clinical and geographical metadata of over 75,000 records in the GISAID COVID-19 database

**DOI:** 10.1101/2020.09.22.20199497

**Authors:** Anjelina Velazquez, Mhealyssah Bustria, Youwen Ouyang, Niema Moshiri

## Abstract

During the SARS-CoV-2 outbreak that caused the coronavirus pandemic it is important now more than ever that scientists and public health officials work side-by-side and use their available resources to track patient information from those that have been affected by the novel coronavirus. The ability to track the disease helps identify possible trends and patterns that can be used by public health officials to make more informed decisions. Tracking data like this may be the key to helping states and countries safely re-open. However, when analyzing large collections of data there is the occurrence of confounding factors such as biases in patient sampling. In this project, a massive collection of COVID-19 data was analyzed, and explored potential biases in patient sampling were explored.

## 1. Introduction

In early 2020, the novel coronavirus, also known as SARS-CoV-2, caused a global pandemic [1]. Due to this, many countries have put their residents under strict public health restrictions and stay-at-home orders. The fast spread of the virus has forced businesses, schools, office buildings and many other public places to shut down and move to remote-work locations. The fast-paced spread of COVID-19 is caused by basic human interactions such as shaking hands and indirectly through objects that have been used or touched by those infected [2]. With this ongoing spread it is important that public health officials have the full visualization of the infection rates, and know the locations where infections continue to rise.

To be able to create visualizations, certain data from the patients being diagnosed with COVID-19 needed to be collected and uploaded by scientists around the world.

The GISAID Initiative was created to enable the faster sharing of data from diseases and viruses such as the novel coronavirus [3]. This initiative proved to be very useful to this research study because of the amount of information that medical facilities around the world were willing to share to this database. The records maintained in the GISAID Database offer biological information about the specimen collected and demographic information about the patient from which the specimen was collected. Databases that encourage data sharing are important because they help researchers from all around the world better understand a global pandemic, and how diseases and viruses are spreading and evolving [3].

The key reason for this study is to observe possible sampling errors that may occur with the collection of data. To combat the COVID-19 pandemic, researchers need to make inferences from data collected from patients. However, for these inferences to be generalizable, researchers need to account for potential confounding factors, such as biases in the patient sampling. In this project, we analyzed a massive collection of COVID-19 data, and we explored potential biases in patient sampling.

## 2. Methods

Patient records were extracted from the GISAID database to form a collection of data on COVID-19 patient records. The process of data collection was performed to be able to identify all of the patient information that has been collected. Python was used to read the collection of data. This data was then analyzed and visualized using the seaborn and matplotlib Python libraries.

### 2.1 Obtaining the collection of data

Data was extracted from the GISAID database. The extracted data was stored in a gzipped JSON file as a dataset of over 75,000 patient specimen records.

### 2.2 Reading the collection of data

Python was used to load the contents of the gzipped file and create a dictionary to hold the patient records.

Each patient record stores an identifier, demographic information about the patient, biological information about the specimen sample that was collected, information about the lab that collected the specimen, and information about the lab that submitted the data to the GISAID database. This study focused on the following attributes:

| Key attributes for this study |  |
| --- | --- |
| Attribute type | Attribute description |
| Accession ID | The record identifier. |
| Gender | The gender of the patient. [4] |
| Location | Where the virus was found. [4] Continent and country are provided, and the names of smaller regions (such as state) may also be provided. |
| Patient age | The age of the host. [4] |
| Collection date | When the sample was collected. [4] |
| Submission Data | When the lab submitted the data for the record. |

Note: Some portions of the attribute descriptions were taken from a source. [4]

While analyzing the data, it was discovered that not all patient records provided the same types of attributes, and the data was not always formatted the same way for a given attribute. Furthermore, some attribute data were not formatted in a way that would be usable in the visualization techniques that we implemented. In general, there were three types of “bad data” that we encountered:

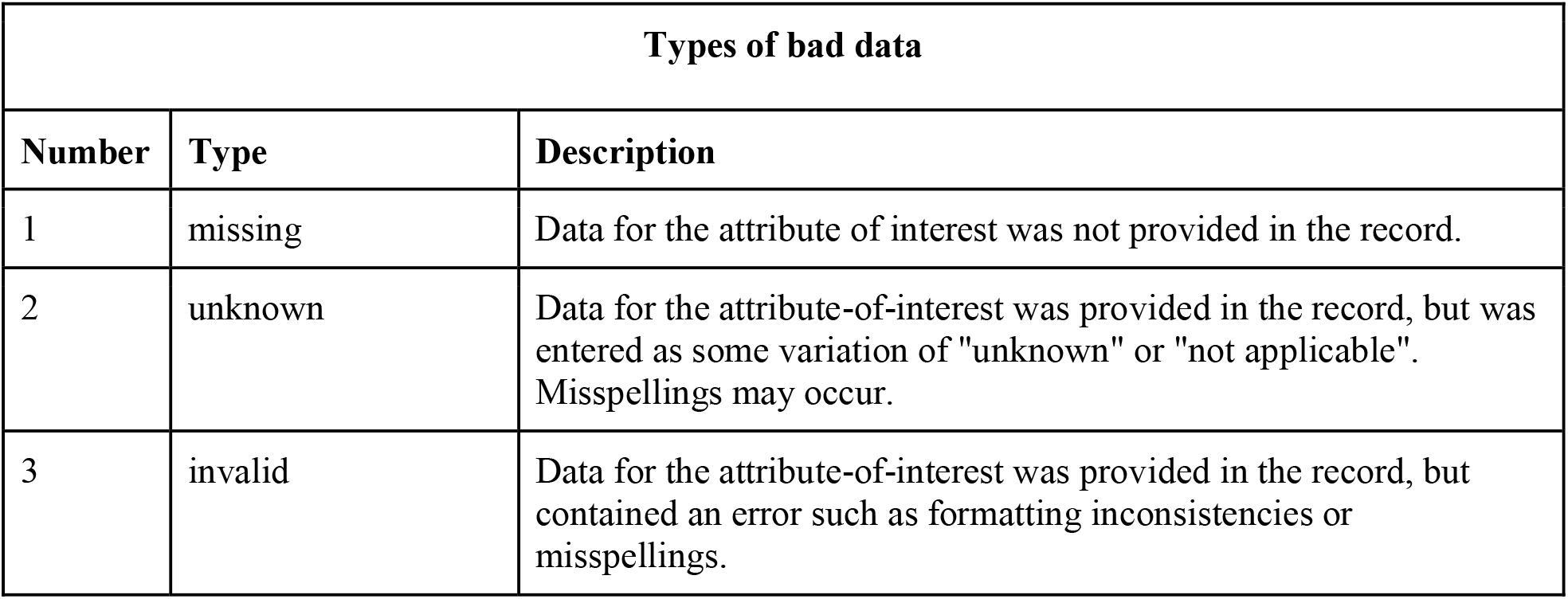

| Types of bad data |  |  |
| --- | --- | --- |
| Number | Type | Description |
| 1 | missing | Data for the attribute of interest was not provided in the record. |
| 2 | unknown | Data for the attribute-of-interest was provided in the record, but was entered as some variation of "unknown" or "not applicable". Misspellings may occur. |
| 3 | invalid | Data for the attribute-of-interest was provided in the record, but contained an error such as formatting inconsistencies or misspellings. |

Python scripts were written to read and display the data that was held in the records. Learning what information was held in the records allowed the identification of data-entry errors that needed to be corrected, and the discovery of information that could potentially uncover interesting trends. To capture as much usable data as possible for analysis and visualization, scripts were written to correct the data-entry errors that were encountered throughout the records. Analyzing the information contained in the dataset was essential for choosing what data to include in the visualizations.

### 2.3 Data Visualization

A variety of figures were generated to visualize trends in the data. Pie charts were generated using the matplotlib Python library. [5] Graphs such as count plots, bar plots, and violin plots were generated using the seaborn Python library. [6]

For many of our charts and graphs, new datasets were created to hold the data required for the visualizations. The type of visualization used to generate each figure was determined by the purpose of the visualization and whether or not frequencies (or “counts”) of a categorical attribute-of-interest were calculated before generating the figure..

| **Types of visualizations used** |  |  |
| --- | --- | --- |
| **Type of visualization** | **Purpose of the visualization** | **Do frequencies (or "counts") need to be calculated before generating the figure?** |
| pie chart | Display the parts that make up a whole. | Yes |
| bar plot | On one axis, display the options for the data values of a categorical variable. On the other axis, display a range for a quantitative variable. Display rectangular bars to represent the relationships between the two variables. | Yes |
| count plot | On one axis, display the options for the data values of a categorical variable. On the other axis, display a range for the frequencies of that categorical variable.. Use rectangular bars to represent the frequency of each data value option. | No |
| violin plot | On one axis, display the options for the data values of a categorical variable. On the other axis, display a range for a quantitative variable. Display violin shapes to represent the kernel density estimation of each data value option. | No |

## 3. Results

### 3.1 Data Collection

At the time of this study, the GISAID database provided 75,571 entries of COVID-19 patient records from around the world. These entries were submitted from countries all around the world, including China, England, Itay, Jamaica, the United States, and many more. During the data collection process many inconsistencies were noticed among the data set. These inconsistencies were due to the fact that the database allows for manually-entered user input instead of automated input. Once inconsistencies like this were fixed, the data collection process gave the study very good results to work with. With over 75,000 records from around the world, getting to see the different continents where the data came from is important. The results below will show the number of records submitted by each continent. The results show the age and gender are among the two highest attributes that were missing throughout the dataset.

### 3.2 Patient Demographics: Country

The following visualizations will show which countries around the world are sharing the most patient records. By yielding data by country, results can be generated to show which gender has a higher infection rate per country. Filtering out the results to only show the countries that have reported 1,000+ records to the database helped to narrow down the results to the countries that need to be focused on the most.

**75**,**571 COVID-19 patient records categorized by country Dates: 2010-3-12 to 2020-5-9**

### 3.2 Dates

One of the attributes that is collected by the database is “Collection date,” which is the day on which the patient’s specimen sample is collected. Another attribute is “Submission Date,” which is the day on which the data for the patient’s specimen sample was submitted to GISAID. Both of these differ greatly from one another, and that it is why it is important to be able to measure the time difference between them both. The results will show how long various countries take to submit the patient data they have collected.

#### 3.2.1 Collection Dates

As previously mentioned, the collection date indicates when the medical facility took the sample of the COVID-19 patient. This is different from the submission date, which indicated when the facility submitted the data to the GISAID Initiative. So results will show the time between submissions was as long as two months at a time. These bar plots below show when COVID-19 specimen samples were collected in China, the United States, England, and Australia.

#### 3.2.2 Number of days between collection of samples and submission of data

In this study the comparison between the submission date against the collection date was done in order to have a better idea of the time elapsed between those dates. The results will show how some countries took longer than others to submit their results to the GISAID Database.

The violin plot shows how quickly data was submitted from China, the United States, England, and Australia. There is a clear comparison between which countries submit their data quicker than other countries. It is important that medical facilities share COVID-19 data as quickly as possible to help public health officials make informed decisions.

### 3.3 Patient Demographics: Location, Gender, and Age

During the data collection process it was noted that for over 60% of the data it was not specified whether the sample was taken from a male or female human. However the rest of the data did have a male or female input. Using this data, plots were made to show the rate of male, female, and unknown gender patients with COVID-19 in different countries.

#### 3.3.1 Location and Gender

#### 3.3.2 Location and Age

In this study relating the age of the infected COVID-19 patients with their location was an important priority. The following plots will show four different countries China, England, Wales, and the USA. In these violin plots the results will show the average age of the people getting infected by the virus. These violin plots are designed to keep public health experts informed about the rising infection rates. Knowing the age of those who are contracting the virus more often than others helps keep the public safety in order The results from the violin plots show how every country has a different average age.

#### 3.3.3 Location, Age, and Gender

The following violin plot will show a combination of three attributes: the Location, Age, and the Gender. Analyzing all three of these allows for the visualization of the different distributions between male and female patients. Female patients will be shown in pink while the male distribution will be shown in blue. The results below show four different violin plots to show the different distribution of four different countries.

## 4. Discussion

### 4.1 Reflections for each chart and graph

The beginning figures provide information about which continents have submitted the most patient records, which were: Europe, North America, and Asia. It is important to keep in mind that inferences can be made about the number of COVID-19 patients in various locations, but the number of records submitted may not necessarily reflect the actual number of people afflicted by the SARS-CoV-2 virus in those locations.

Further into the results section there were plots that provide information about the data for patient gender, and the patient age that was found in the dataset. When the gender and the age attribute was taken throughout the dataset, the unknown amount is significantly higher than the amount of data that was properly submitted. Due to this bad data, almost 70% of the records in the dataset were excluded when analyzing patient gender and patient age. Due to this high percentage of unknown data a bar plot was provided to inform about how often bad data was encountered in this study. The results showed gender and patient age were two of the highest records in the dataset that had missing, unknown, or invalid data. Also, note that one record may be missing multiple attributes at a time.

Also in the results section many plots provide information about data-submitting practices and COVID-19 rates in various countries. In the time range specified in figure 6, ten countries have submitted over 1,000 patient records. The result showed that England, USA, Wales, Scotland, and Australia have submitted the most patient records. With so many records a plot was provided to identify information about when the specimen samples were collected in various countries. Results showed the country of China collected the majority of their samples in the earlier months of the year 2020. The results also suggest that China had an earlier pandemic than the rest of the countries. China has the highest rate of “before 2020” collection. Inference can be made that most of the collections made in the United States, England and Australia were done during the early months of spring and all throughout the summer. The plot also shows that the United States started collecting samples later than China. These plots show that all three countries had a soaring amount of collections during the same months. This suggests that the United States, England, and Australia experienced the worst of the COVID-19 pandemic at the same time.

**Figure 1:**
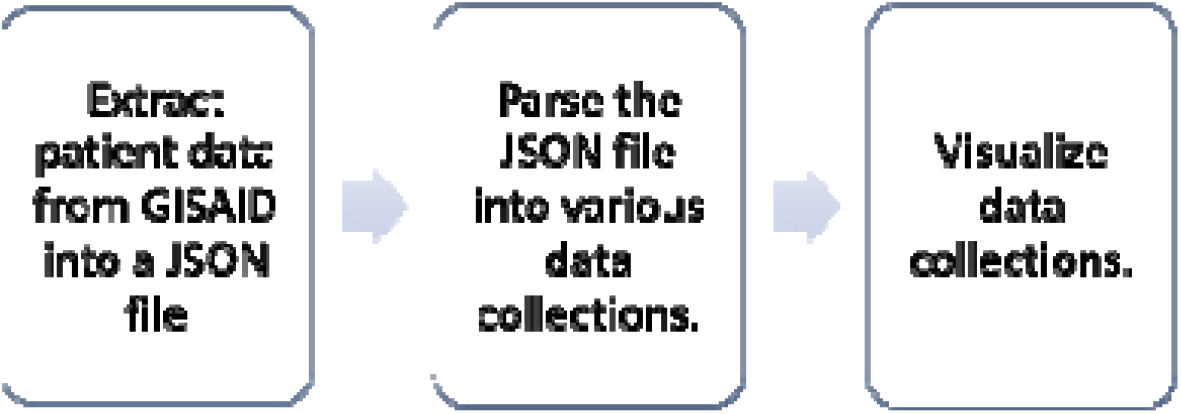
The process of obtaining over 75,000 patient records and generating visualizations.

**Figure 2:**
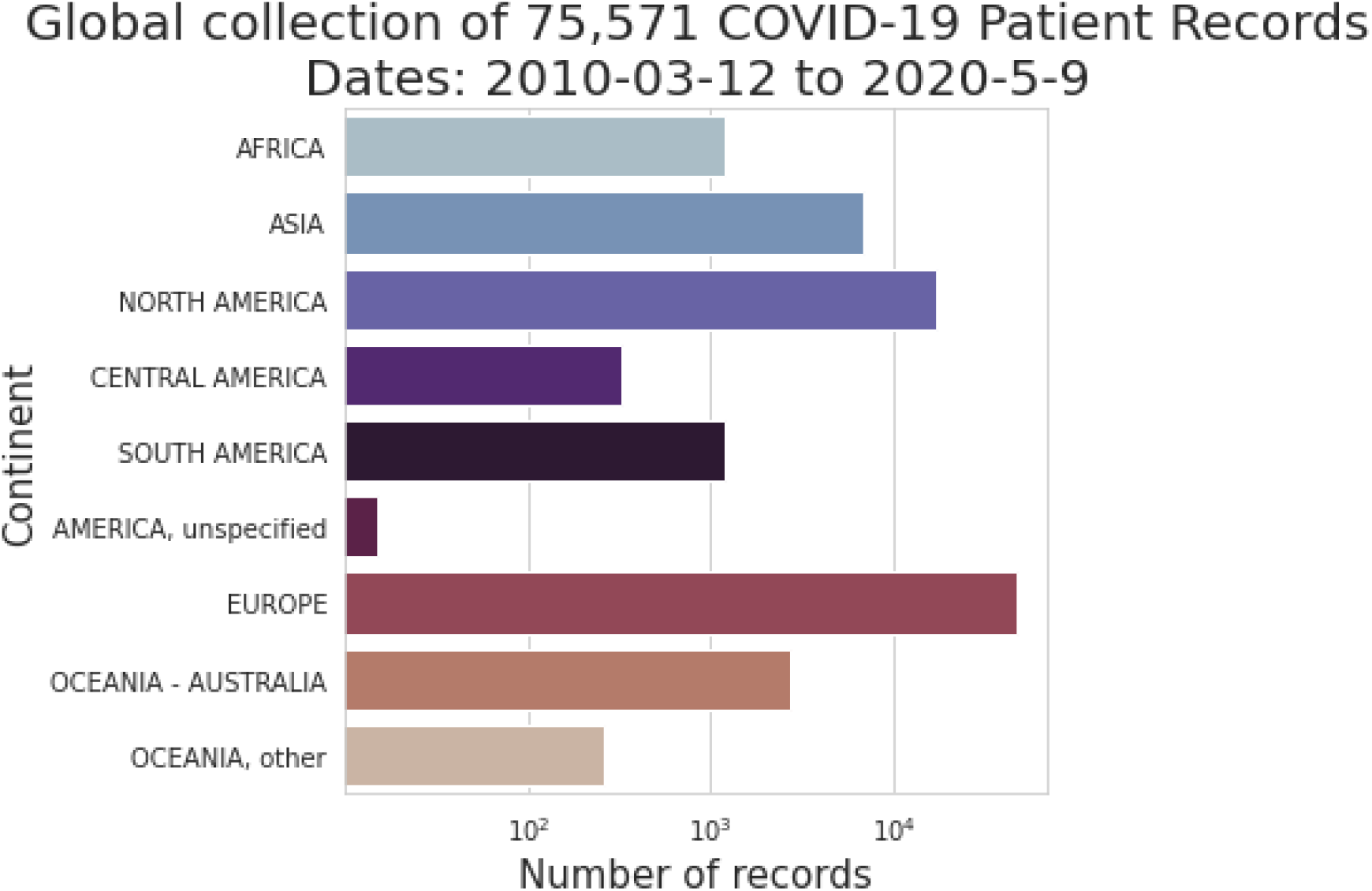
The database provided data for 75,571 patient records from around the world. This bar plot shows the number of patient records submitted from each continent that was found in the dataset.

**Figure 3:**
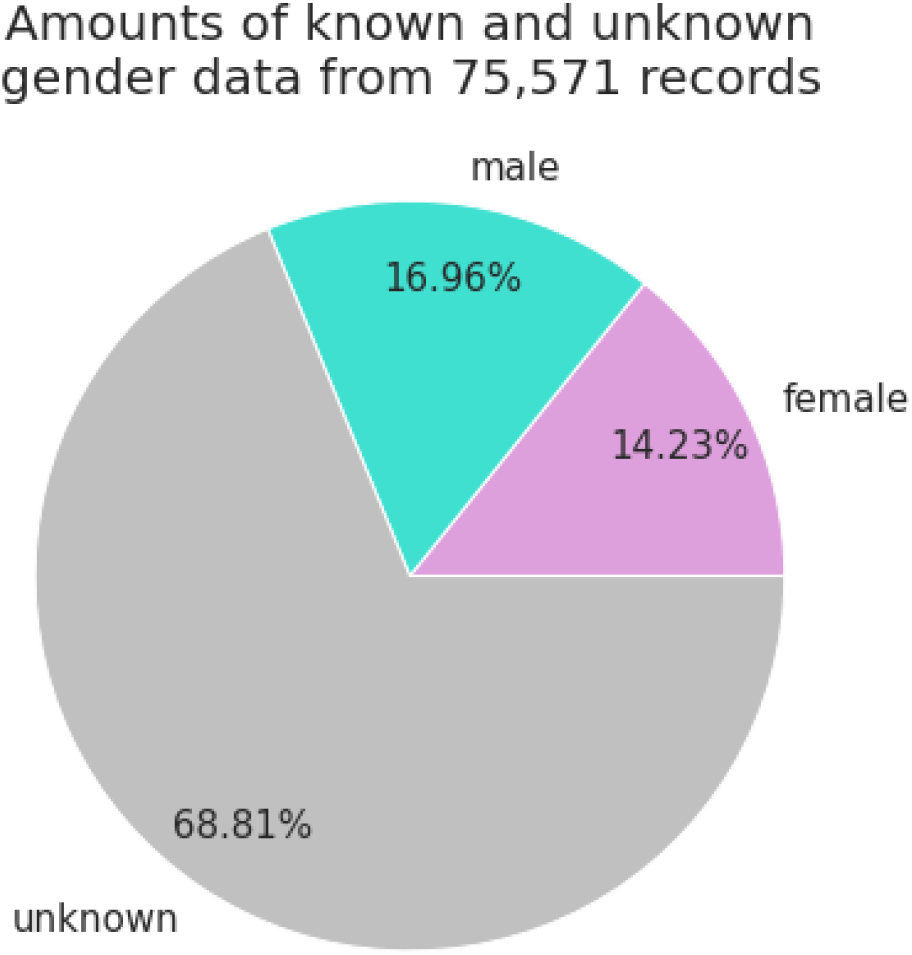
This pie chart shows the amounts of known and unknown gender data in the dataset. Some records provided gender as either female or male, and the others provided gender as unknown. Female patients are in pink, male patients are in blue, and the unknown is in gray.

**Figure 4:**
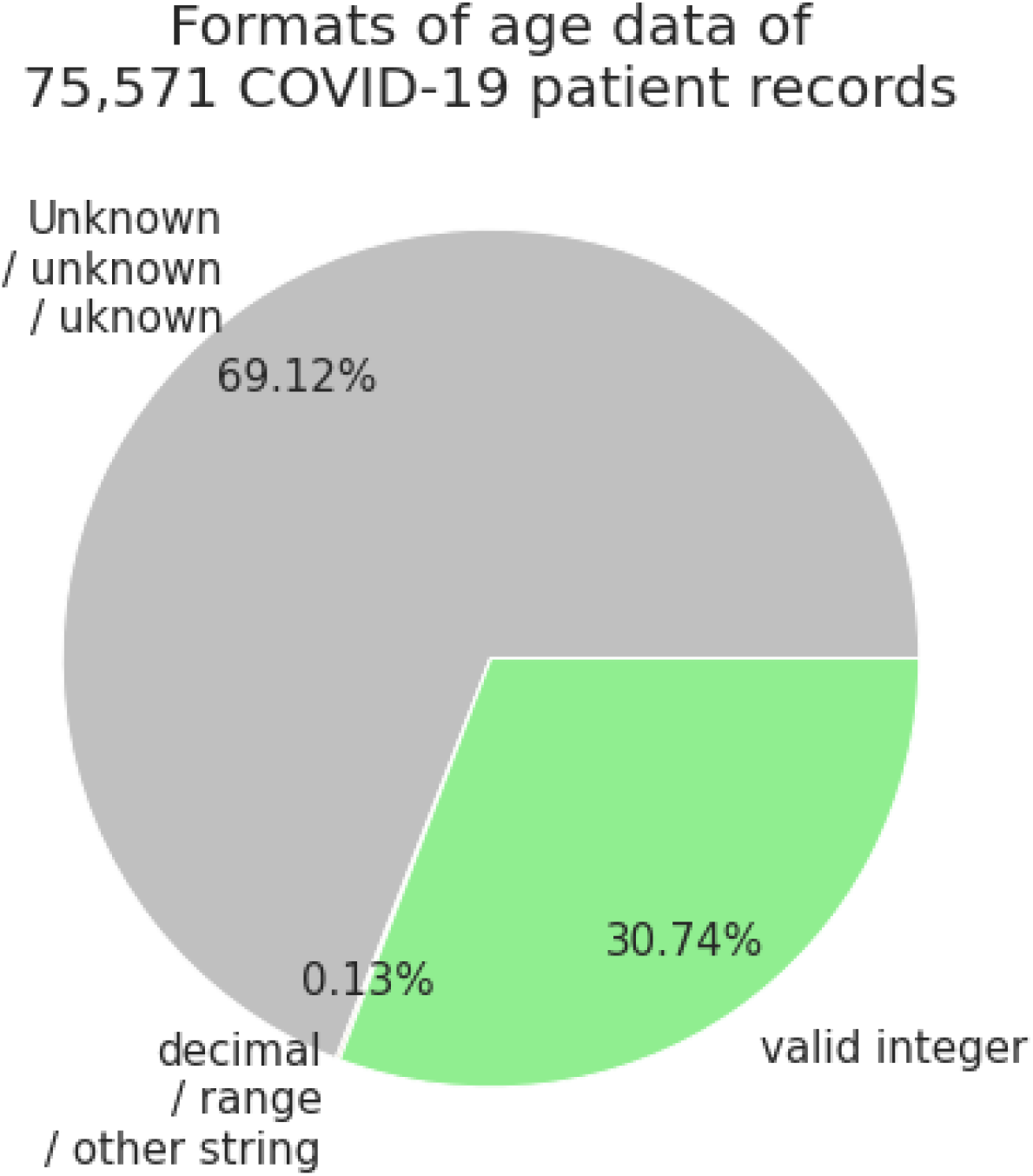
This pie chart is a categorization of the patient age data that was found throughout the dataset. Ages were provided as either an integer, a string indicating “unknown”, or as other non-standard strings. The results show that only about 30% of the age data was provided.

**Figure 5:**
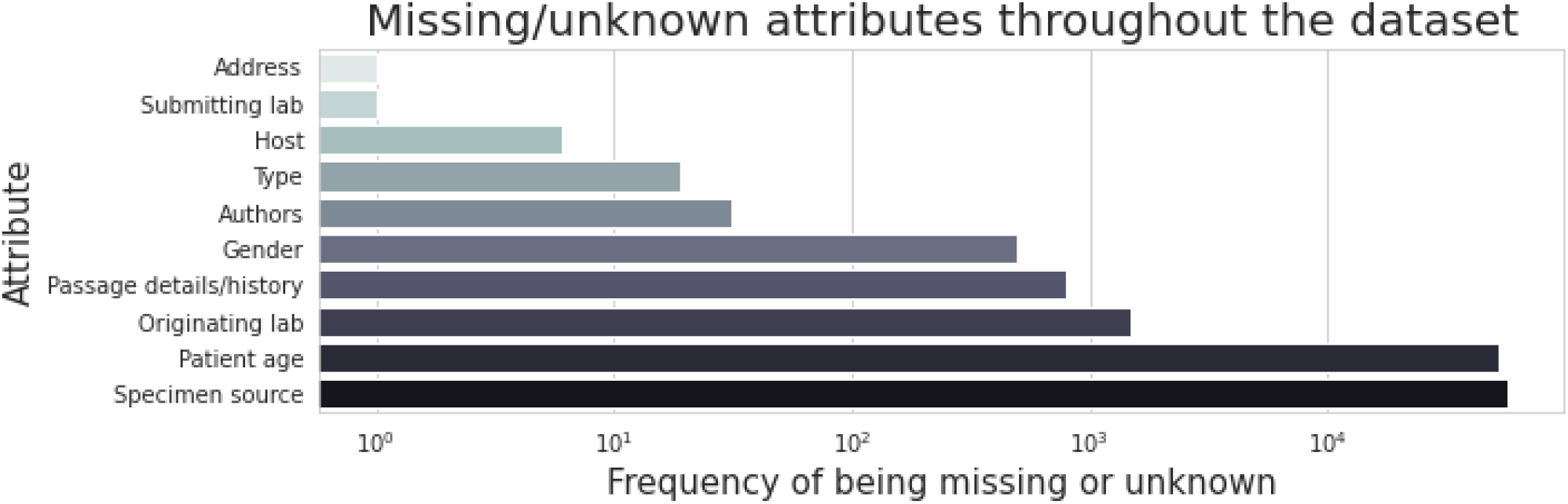
This bar plot shows how often data was missing or unknown in the dataset. The y-axis shows various attributes, and the x-axis represents how often bad data was encountered for these attributes.

**Figure 6:**
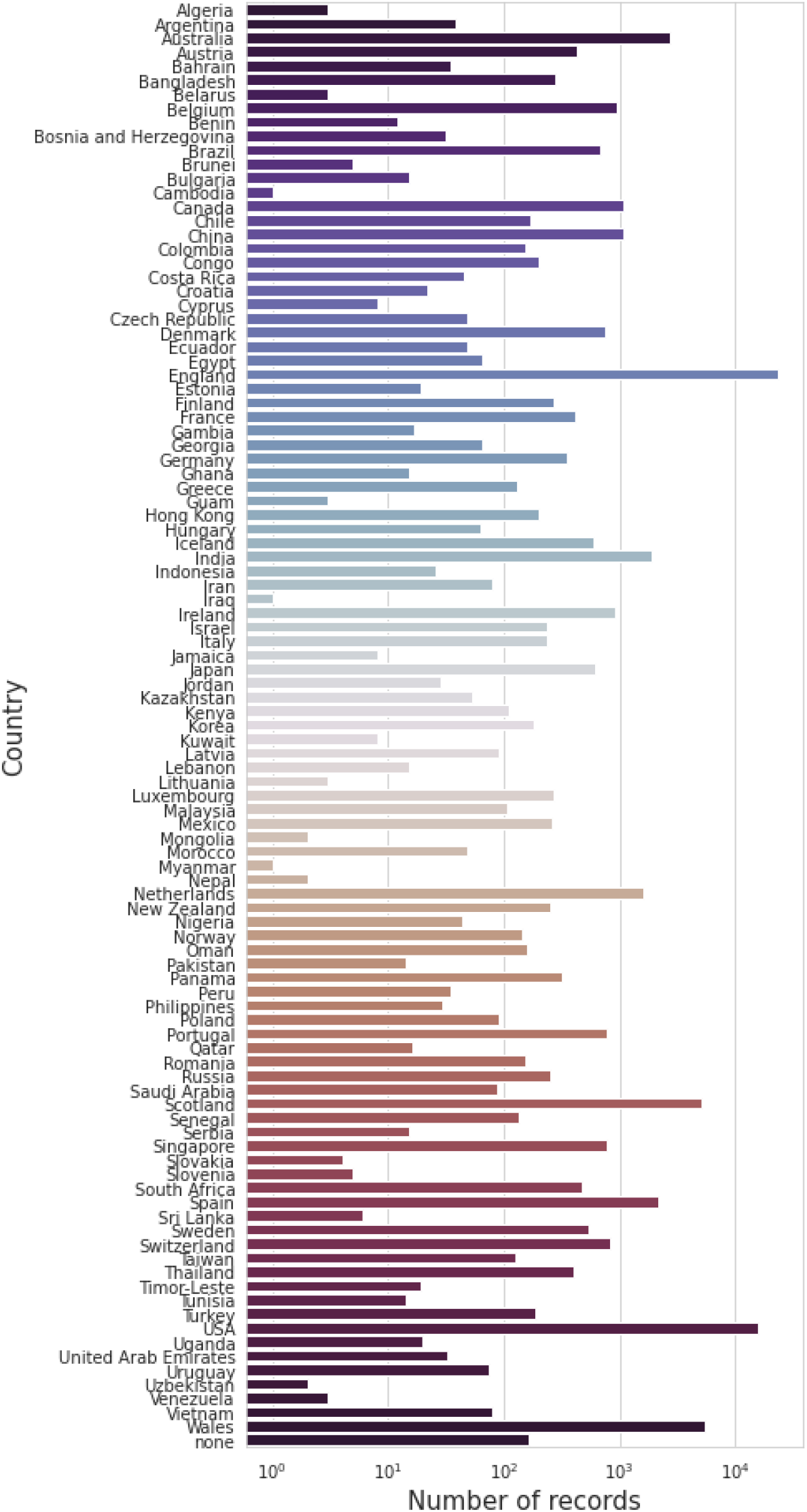
This count plot shows the number of COVID-19 patient records submitted by each country. On the y-axis is the country’s name and on the x-axis is the number of records.

**Figure 7:**
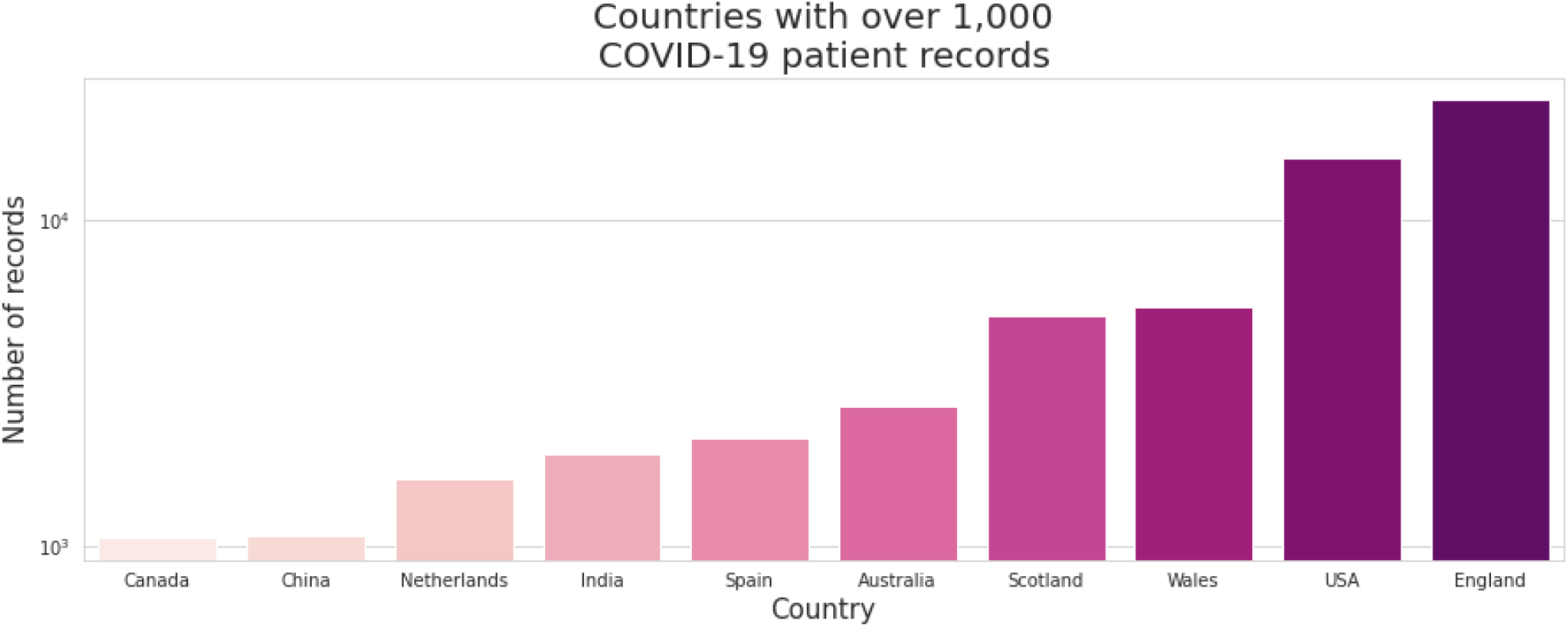
This bar plot shows the countries that have submitted over 1,000 records. The number of records in log-scale are on the y-axis and the name of the country on the x-axis.

**Figure 8 (a):**
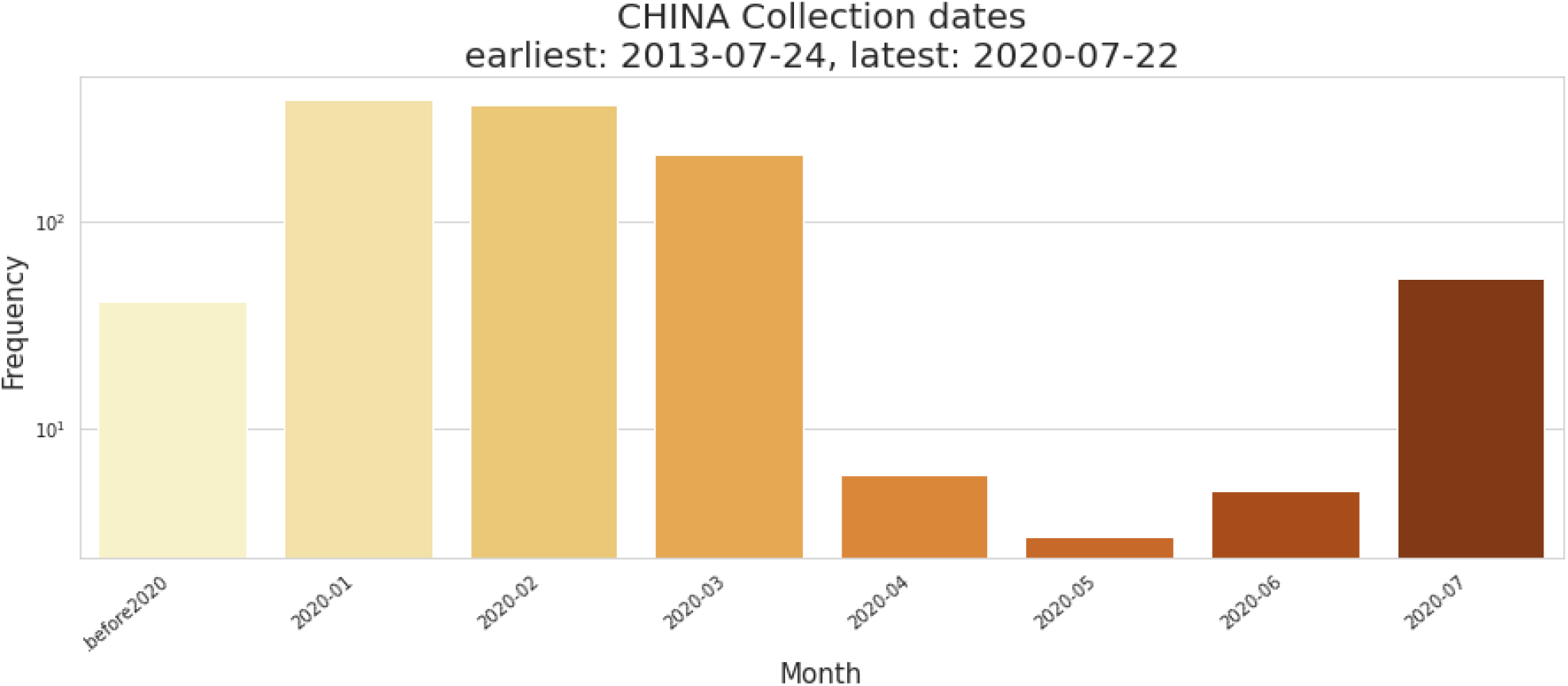
This bar plot visualizes the collection dates that were attached to each of the patient records that were submitted from China. On the y-axis the frequency is listed (number of records) and on the x-axis we have the date intervals as monthly. This means that between the months of January and March, China submitted over 100 records per month.

**Figure 8 (b):**
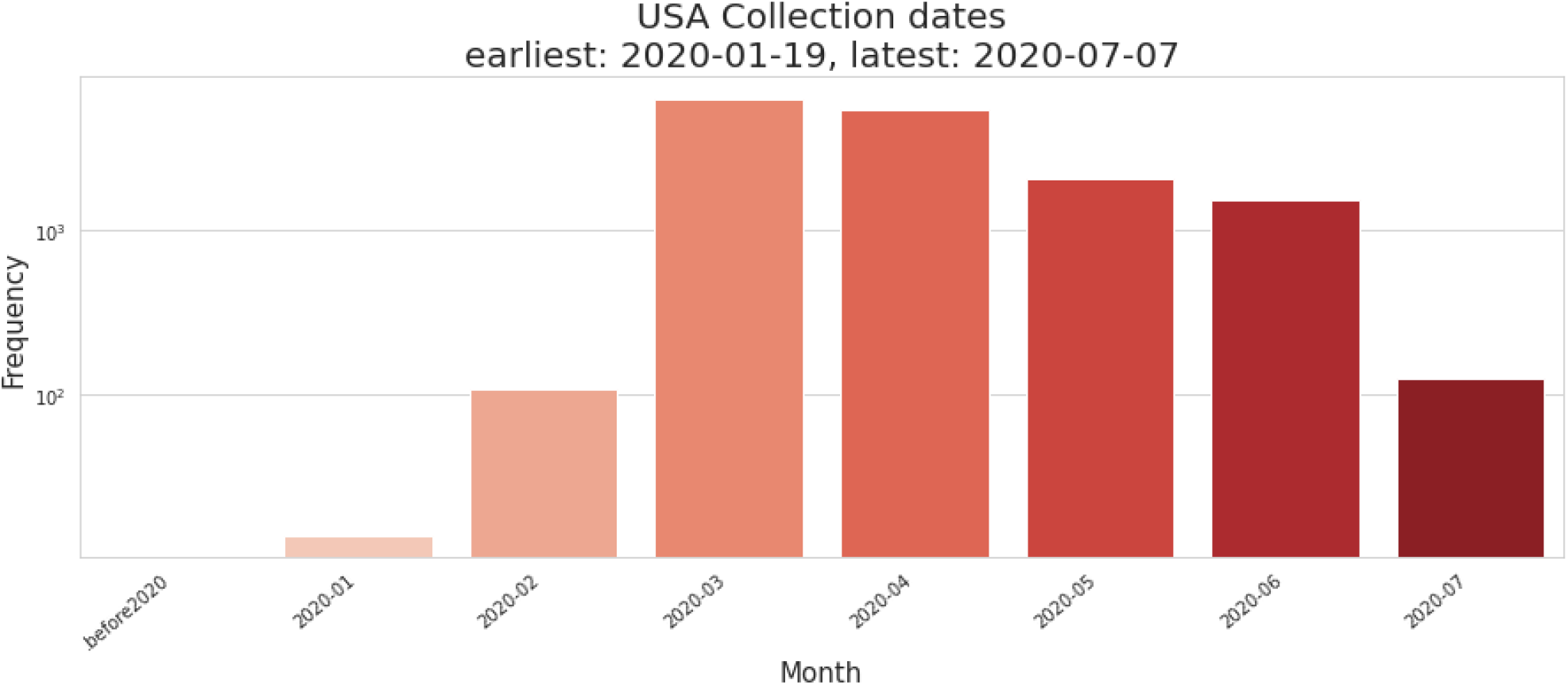
This bar plot visualizes the collection dates that were attached to each of the patient records that were submitted from the United States. On the y-axis the frequency is listed (number of records) and on the x-axis we have the date intervals as monthly. The results show that between the months of March and June, the USA submitted over 1,000 records.

**Figure 8 (c):**
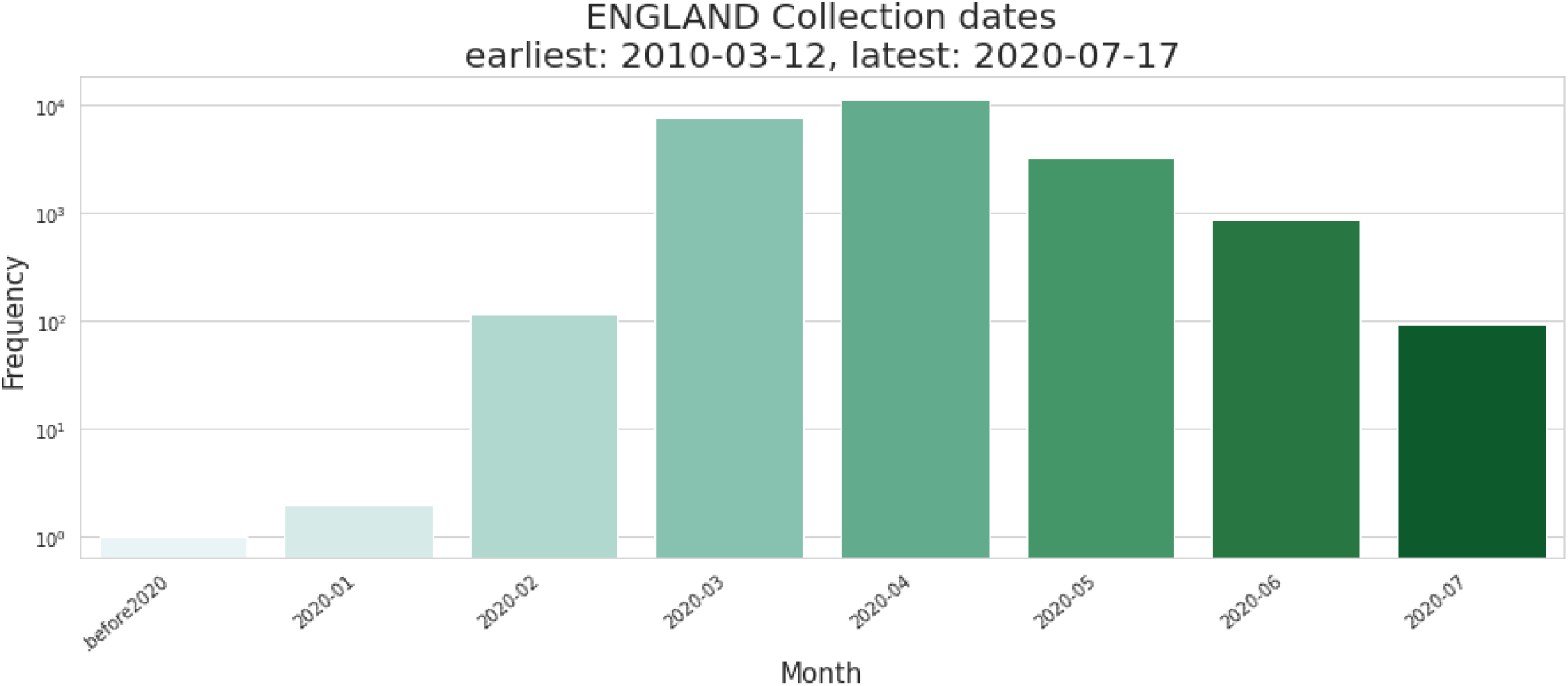
This bar plot visualizes the collection dates that were attached to each of the patient records that were submitted from England. On the y-axis the frequency is listed (number of records) and on the x-axis we have the date intervals as monthly. Between the months of March and May, England submitted over 1,000 records.

**Figure 8 (d):**
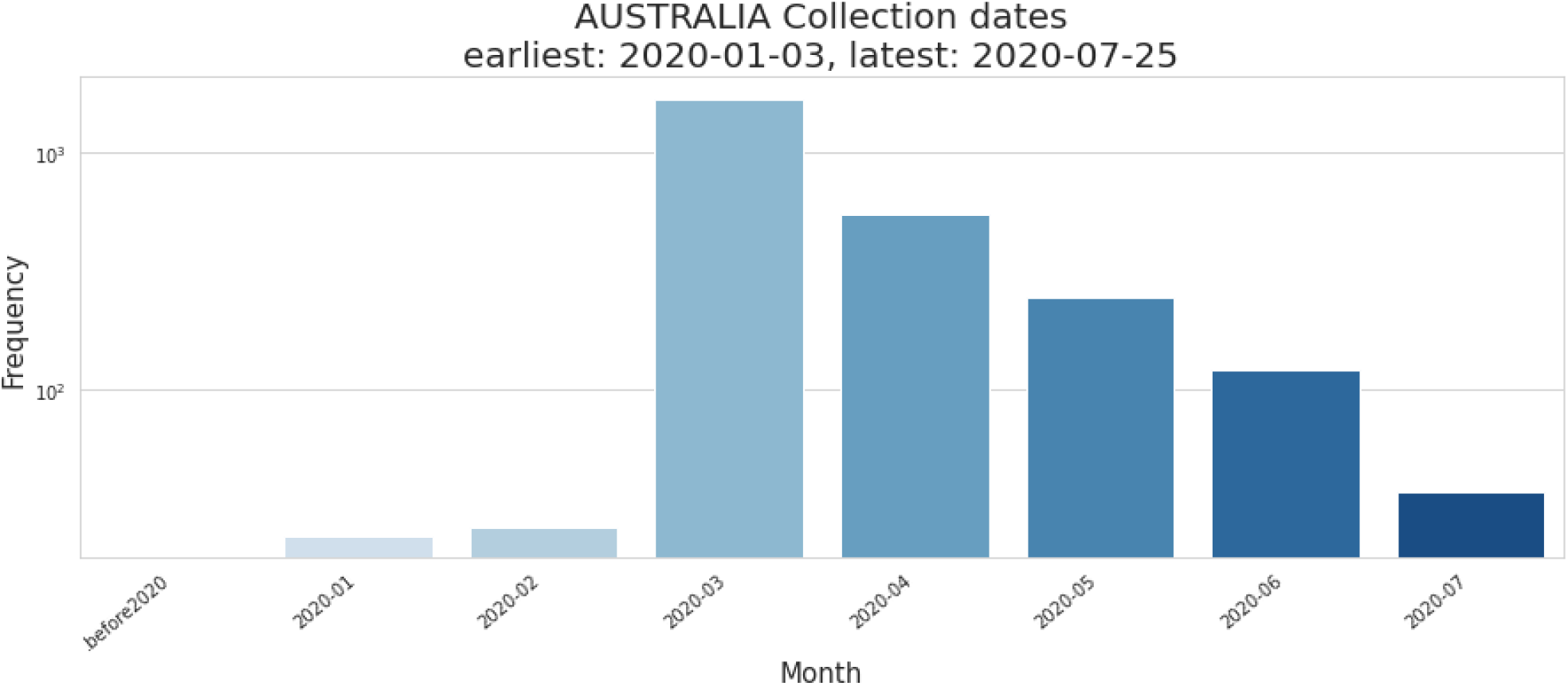
This bar plot visualizes the collection dates in Australia. On the y-axis the frequency is listed (number of records) and on the x-axis we have the date intervals as monthly. The results show that, in the month of March, over 1,000 were reported.

**Figure 9:**
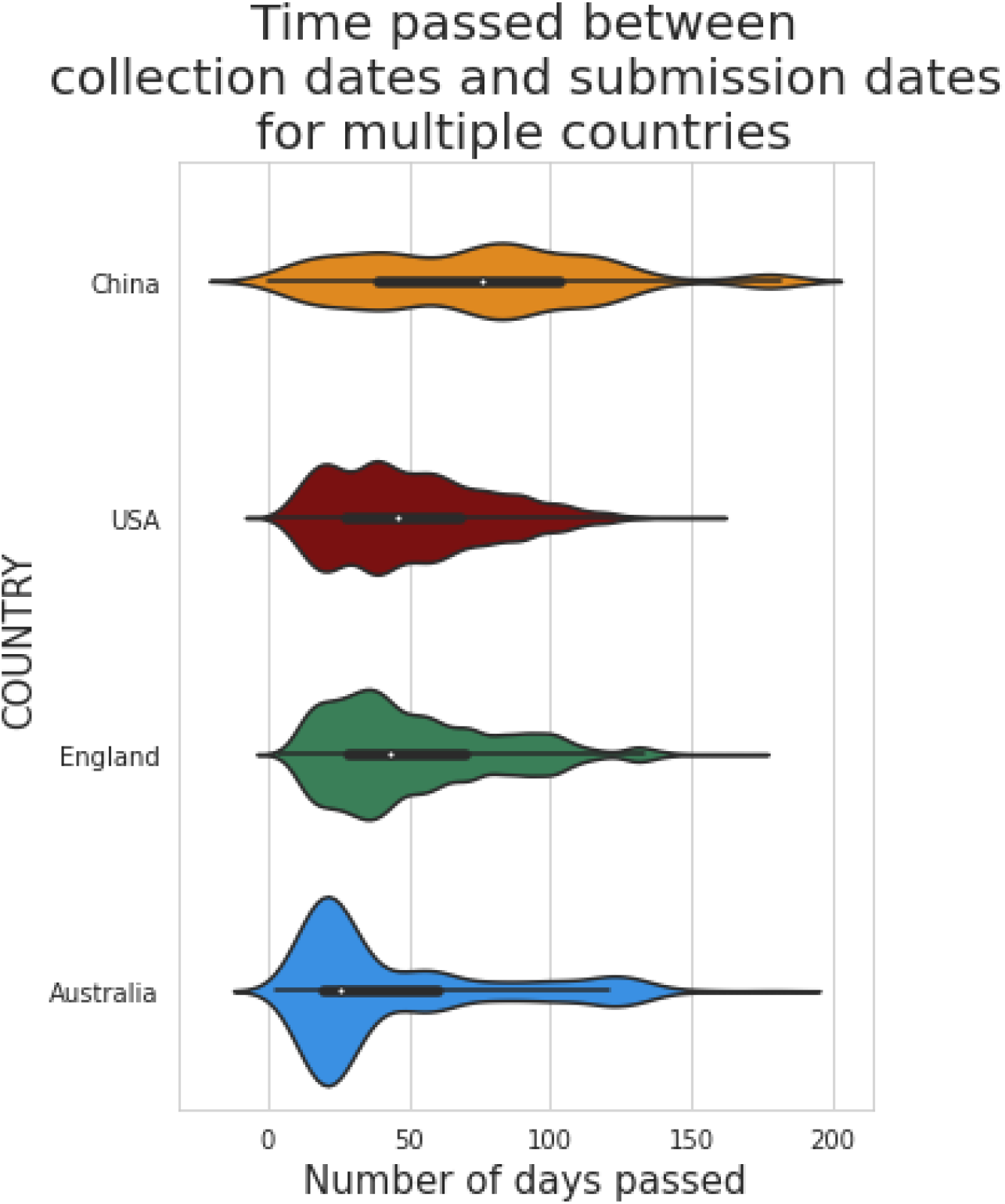
This violin plot shows the kernel density estimates of the time elapsed to submit data after a sample had been collected in China, the United States, England, and Australia. On the y-axis the country is listed and on the x-axis the days are listed. The results show the USA, England, and Australia submitted records in under fifty days.

**Figure 10:**
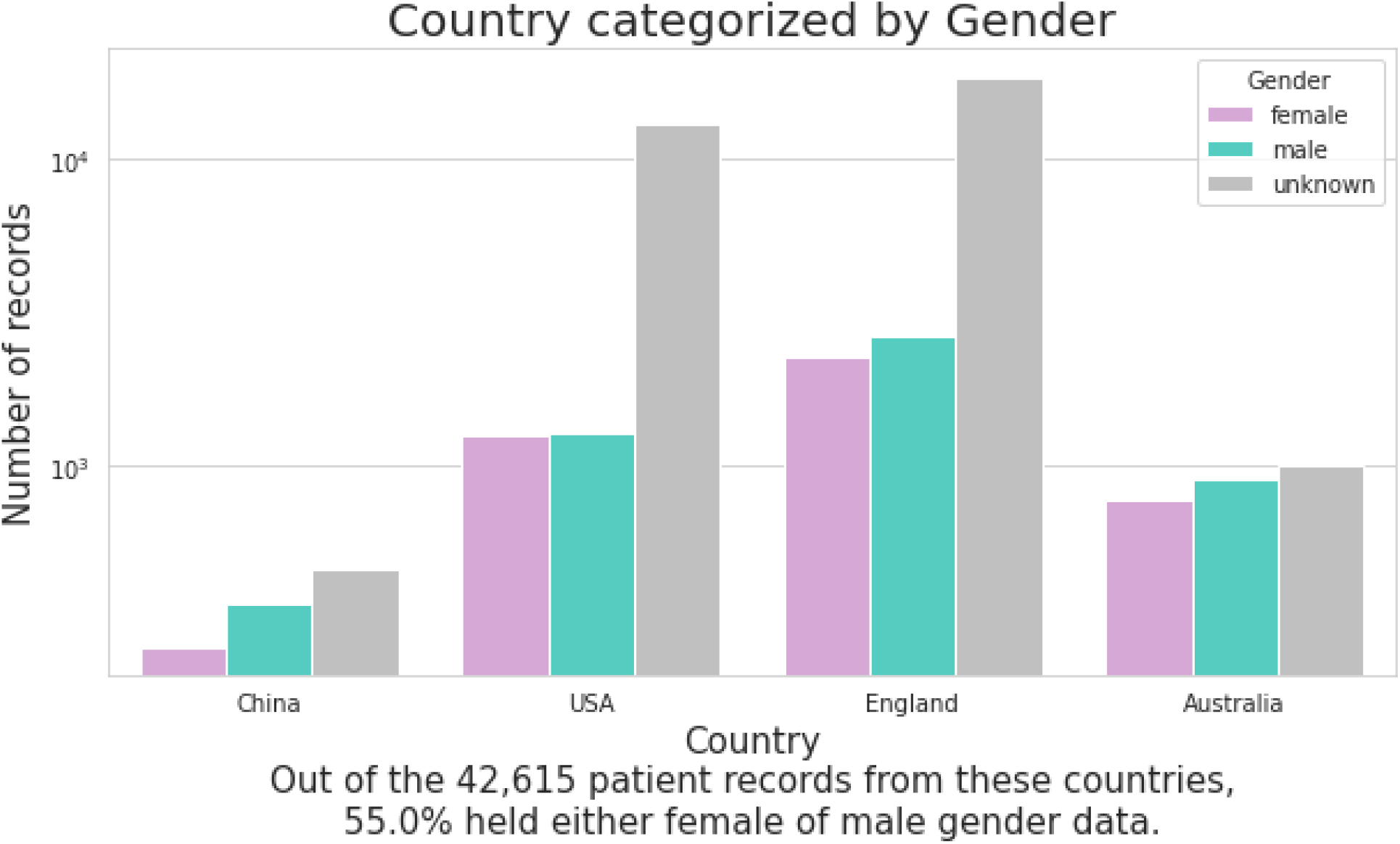
This count plot shows the frequencies of female and male COVID-19 patients in the records from China, the United States, England, and Australia. The number of records in log scale are on the y-axis and the country is on the x-axis. The color pink represents all the female patients, the color blue is all male patients, and gray is all the patients that did not have a gender for. The results show that England and the USA are imputing the most unknown data for gender.

**Fig 11:**
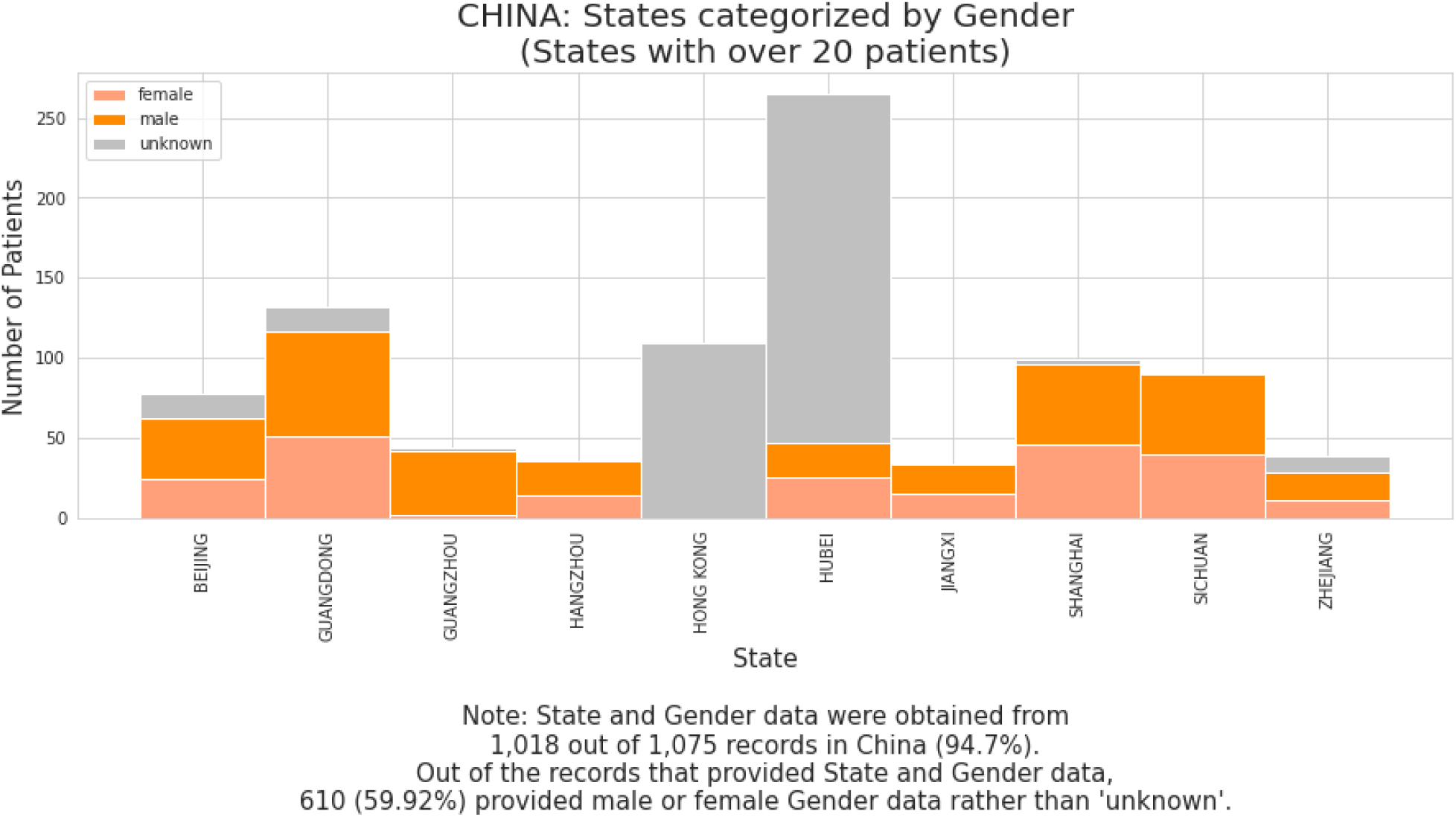
These stacked bar plots show what portion of each state in China has records for patients of female, male, or unknown gender. On the y-axis the number of patients is listed and on the state names are on the x-axis. The results show that a majority of unknown records come from Hubei.

**Fig 11 (b):**
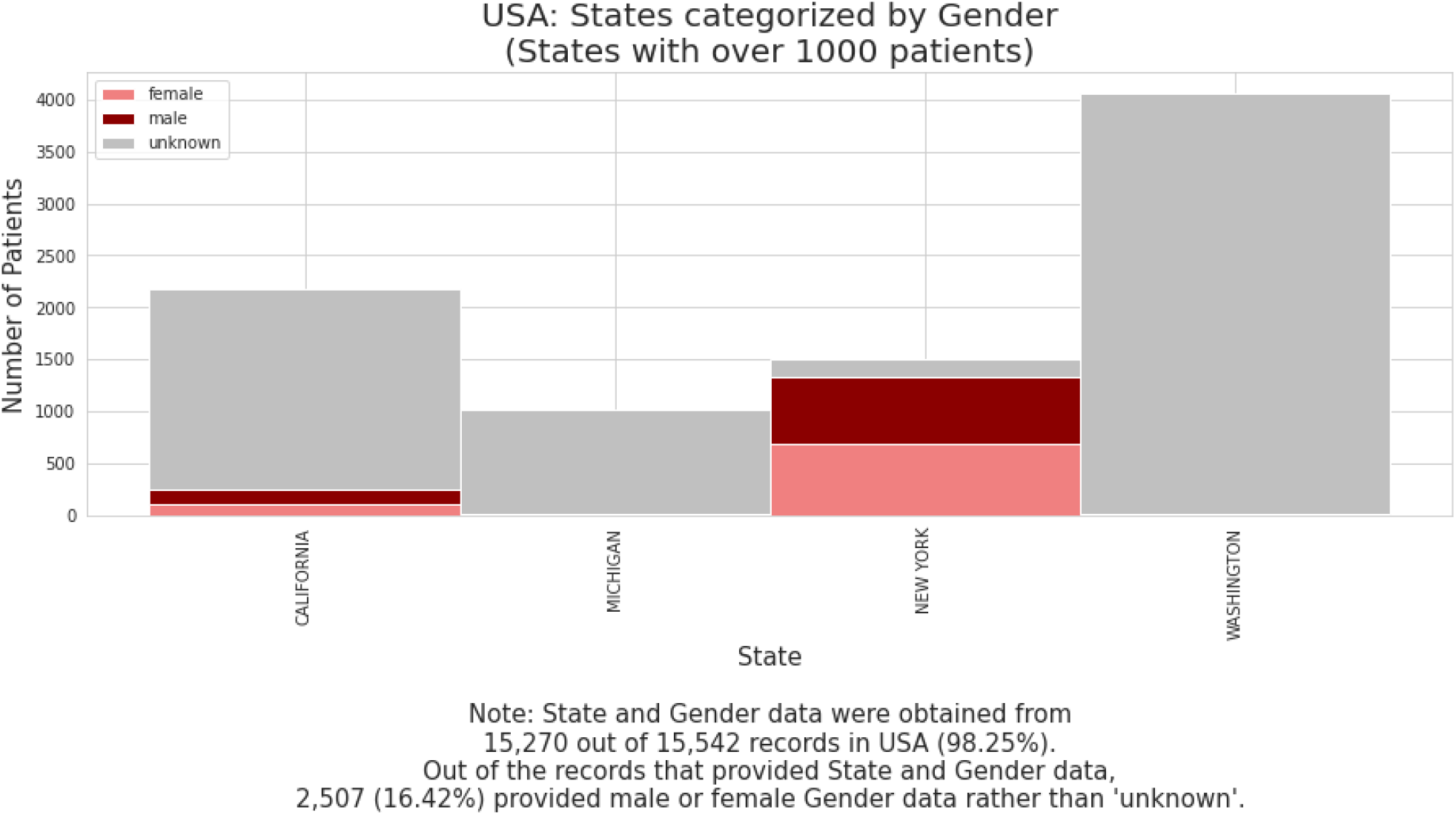
These stacked bar plots show what portion of each state in the United States has records for patients of female, male, or unknown gender. On the y-axis the number of patients is listed and on the state names are on the x-axis. This plot shows that Michigan and Washington have submitted only unknown Gender data.

**Fig 11 (c):**
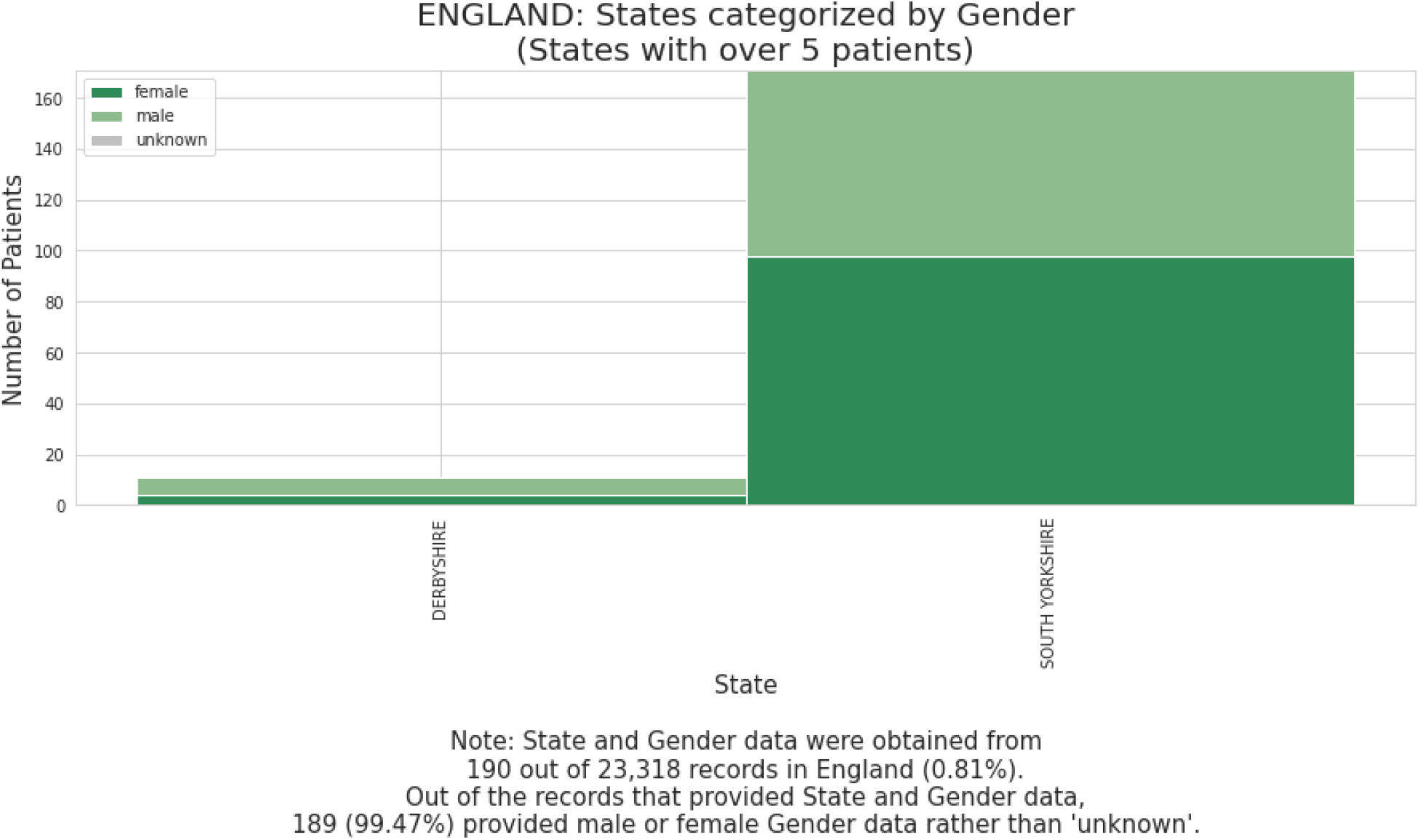
These stacked bar plots show what portion of each state in England has records for patients of female, male, or unknown gender. On the y-axis the number of patients is listed and on the state names are on the x-axis. The plot shows that these states have not produced any unknown data.

**Fig 11 (d):**
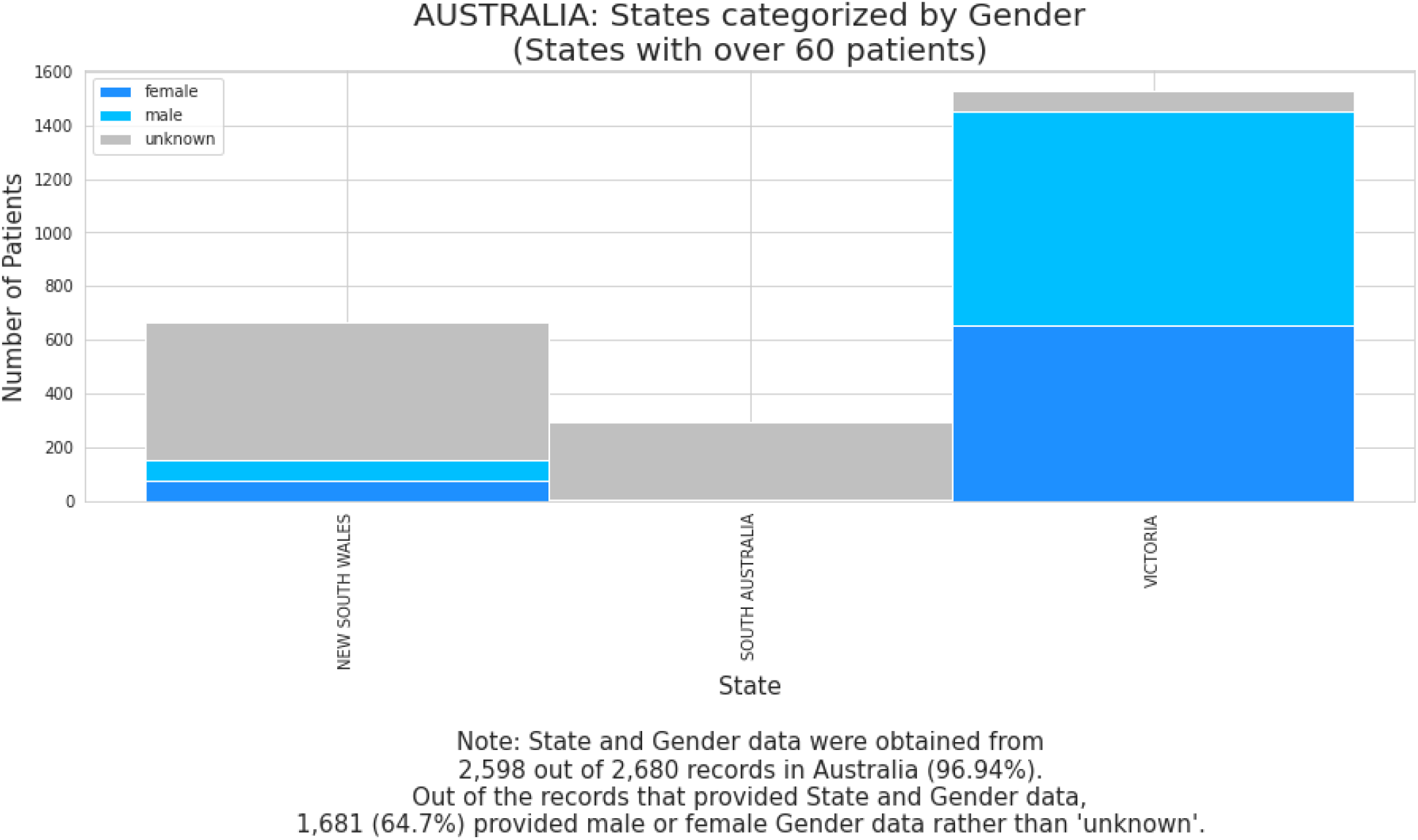
These stacked bar plots show what portion of each state in Australia has records for patients of female, male, or unknown gender. On the y-axis the number of patients is listed and on the state names are on the x-axis. This plot fromAustralia shows that the state Victoria has the most amount of data compared to the other two states.

**Figure 11:**
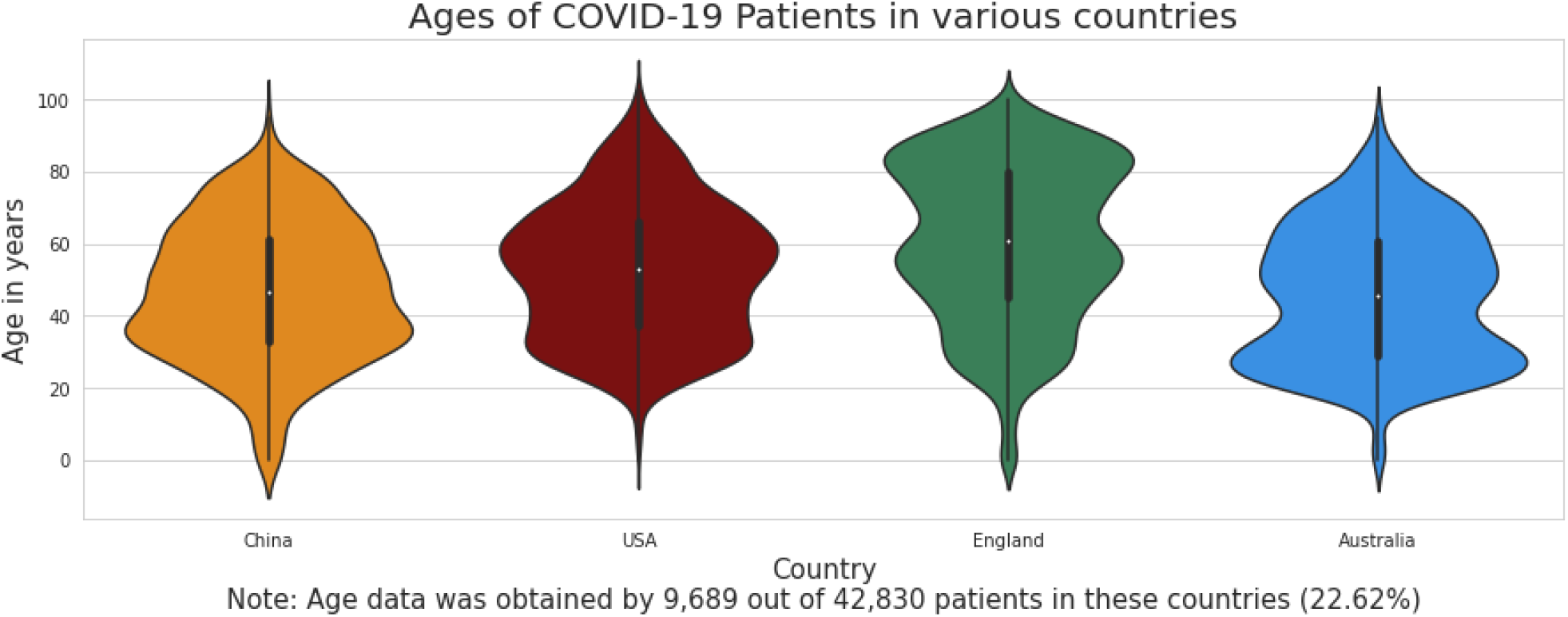
Age (in years) are on the y-axis, and countries are on the x-axis. This violin plot shows the probability distributions for the ages of COVID-patients in China, England, the United States, and Australia. The results show that China, Australia, and the USA have a median age of over 40 years old. While England shows a higher median age at around 60 years old.

**Fig 12:**
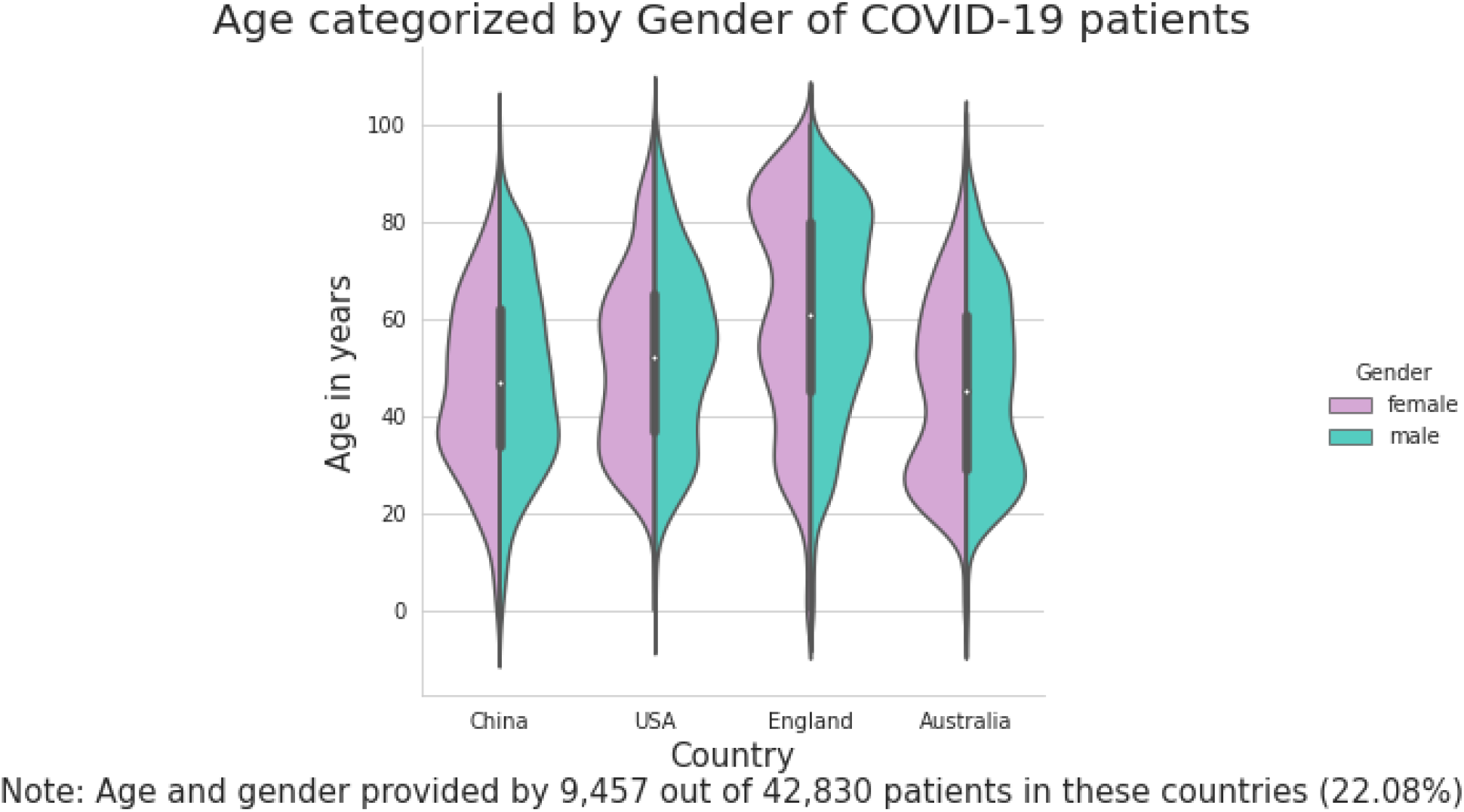
Once again the age in years is on the y-axis and the countries are on the x-axis. The violin plot has been split to show all female records in pink and the male records in blue. This split violin plot shows the distribution of patient ages for female and male patients in China, the United States, England, and Australia. In China and Australia,the distributions for male and female patient ages are even. In the USA, there is a heavier distribution of male patients above 50 years old. And In England, there is a heavier distribution of females under 40 years old.

Plots were also made to provide information about how much time has passed between sample collection and data submission in various countries. China generally took between 50 and 100 days (2-3 months) to submit COVID-19 patient data after taking a specimen sample, which is slower than what was observed in the USA (0-2 months), England (0-2 months), and Australia (0-2 months). Among these four countries, Australia is shown to be likely to submit patient data within 1 month of taking a specimen sample.

The violin plots that were made provide information about the gender data provided by the patient records in China, the USA, England, and Australia. In each of these four countries, the number of patient data increases from female to male to unknown. That is, the number of male patient data is higher than the number of female patient data, and the number of patient data of unknown gender is higher than the numbers of patient data for both females and males. In China and Australia, the number of unknown-gender patients is higher but relatively similar to the numbers of female and male gender patients in the respective countries. In the USA and England, the number of unknown-gender patients is much higher than the numbers of female and male gender patients in those countries.

Additionally violin plots were also used to provide information about patient ages in various countries. China, the USA, and Australia have a median age of over 40 years old. England shows a higher median age: around 60 years old.In a different plot the results show that in China and Australia, the distributions for male and female patient ages are even. In the USA, there is a heavier distribution of male patients above 50 years old. In England, there is a heavier distribution of females under 40 years old.

### 4.2 The impact of bad data

Missing, unknown, and invalid data (“bad data”) resulted in smaller sample sizes, which could limit the generalizability of inferences made from data analysis. As discussed in Section 4.1, almost 70% of patient records had bad data for gender and age.

Other than the issues with gender and age data discussed in Section 4.1, location data was also difficult to read and to accurately discern usable data. Regarding the location data provided in the records, the formatting was inconsistent, and it is also common to encounter inconsistent naming conventions, and even typos.

To prevent the inclusion of bad data when performing analyses, bad data had to be handled on a case-by-case basis. Error-detection methods were implemented throughout the study. When data for an attribute-of-interest was missing in a record, that record had to be excluded from the analysis. When formatting or spelling errors were encountered, either data-exclusion or data-correction was performed, depending on whether or not the correction could be confidently-assumed from the bad data.

### 4.3 Possible improvements for data entry practices

If a dataset is not cleanly-structured, the computational analysis process is adversely affected. Some potential improvements to data entry practices are improving manual data entry and using automated data capture instead of manual data entry. If manual data entry is used, the process can be improved by the use of dropdown menus (rather than free text) and data validation to ensure that all information is formatted correctly. If automated data capture (such as reading data directly from a file) is used instead of manual data entry, the precision and correctness of the data would be greatly improved, along with increasing the efficiency of the data entry process.

Although error-detection and error-correction methods were used during this study, there are risks involved with handling data this way. There is a high chance that errors may go undetected and will not be excluded in the dataset, which would cause bad data to be included in the analysis. If data is excluded, the sample size to make inferences from is smaller. Data-correction is preferred over data-exclusion, if possible, but is still not risk-free. If error-correction is used, there is risk involved with choosing to assume what the corrected value of the data is.

Along with the risks involved, there were also impacts on the implementation of the code used in this study. As new errors were discovered throughout the study, new code was written to handle those errors. Many errors were handled using very specific code rather than being generalizable. For example, specific code was written to handle individual abbreviations, misspellings, and other alternative naming conventions. One tool that helped improve the efficiency of reading and filtering the data was a predefined list of countries. The data-handling process could potentially be further improved by utilizing predefined lists, such as lists of all cities in a country.

## Data Availability

All raw data is available on the GISAID database. The Jupyter notebook written to perform the data analysis can be found here: https://colab.research.google.com/drive/12KULPYkIQsxc-v8ayZcIEtf43tOxZfkh?usp=sharing

https://www.gisaid.org/

https://colab.research.google.com/drive/12KULPYkIQsxc-v8ayZcIEtf43tOxZfkh?usp=sharing

## 5. Data Availability

The Google Colab Notebook for this project can be found at: https://colab.research.google.com/drive/12KULPYkIQsxc-v8ayZcIEtf43tOxZfkh?usp=sharing

## 6. Acknowledgements

This work was supported by NSF grant NSF-2028040 to N.M., the Google Cloud Platform (GCP) Research Credits Program, and the UC San Diego Department of Biomedical Informatics (DBMI) Short Term Training Program (STTP).

